# Local Aromatase Activity Alterations in Breast Cancer Tissues: A Potential Way of Decision Support for Clinicians

**DOI:** 10.1101/2020.06.04.20122101

**Authors:** Mete Bora Tuzuner, Tulin Ozturk, Sennur Ilvan, Hande Turna, Turkan Yurdun, Hulya Yilmaz-Aydogan, Oguz Ozturk

**Affiliations:** Research and Development Center, Acibadem Labmed Medical Laboratories Içerenkoy Mah, Kerem Aydinlar Kampusu, Kayisdagi Cd. No:32, 34752 Atasehir/Istanbul Turkey; Department of Pathology, Cerrahpasa Medical School, Istanbul University Cerrahpasa Cerrahpaşa Mahallesi, Koca Mustafapaşa Cd. No:53 D:No:53, 34096 Fatih/İstanbul Turkey; Department of Internal Medicine, Section of Medical Oncology, Cerrahpasa Medical School, Istanbul University Cerrahpasa Cerrahpaşa Mahallesi, Koca Mustafapa İa Cd. No:53 D:No:53, 34096 Fatih/İstanbul Turkey; Department of Pharmaceutical Toxicology, Faculty of Pharmacy, Marmara University Tıbbiye cad. No.49, 34668 Haydarpaşa /İstanbul; Department of Molecular Medicine, Aziz Sancar Institute of Experimental Medicine, Istanbul University, Vakıf Gureba Cad. Çapa Kampüsü Şehremini - 34390 Fatih / İstanbul Turkey

**Keywords:** Breast cancer, tumor microenvironment, aromatase, estrogens, mass spectrometry

## Abstract

**Background and aims:** It is becoming evident that local estrogen exposure is important in postmenopausal breast cancer patients. The microenvironment is established by breast stromal cells based on communication with tumor cells that is essential to cancer development, invasion, and metastasis. Here we investigated aromatase activity levels in both tumor and matched stromal tissues by showing their impact on the manufacturing of local estrogen and tumor progression in cases of invasive ductal carcinoma (IDC).

**Methods:** Tumor (T) and tumor-associated stroma (TAS) neighboring tissues were acquired from each postmenopausal patient, diagnosed with IDC, and categorized as luminal A (n = 20). The control group was formed from tumor-free breast tissue samples (N, n = 12). A microsomal-based technique was created to compare breast tissue aromatase activities using liquid chromatography – mass spectrometry.

**Findings:** We observed that the TAS tissues have the highest aromatase activities (p < 0.05). High progesterone receptor (PR) intensity levels were found to be decreasing the activity level in these tissues significantly (p< 0.05). Tumor tissue specific aromatase activity levels of postmenopausal patients’ were tend to be lower compared to healthy premenopausal subjects’ (3 fold, p< 0.001). In addition low activity in tumor tissues were associated with low grade and late stage cancers.

**Conclusions:** Early detection and personalized therapy is essential for postmenopausal breast cancer patients. Together, our inhouse tandem mass spectrometry technique has the potential for further development and standardization for the measurement of aromatase activity and may assist clinicians decide on therapy policies for postmenopausal IDC patients which could be an invaluable asset for precise and specific evaluation.

## 1. Introduction

The second most prevalent cancer in the globe is breast cancer, and by far the most prevalent cancer among females [1]. Almost 75% of the cases are hormone-dependent and estrogen receptor (ER) positive [2]. Estrogens act as intracrine and paracrine factors (rather than a circulatory hormone) in the breast tissue of postmenopausal women [3,4].

Ovaries significantly cease to supply estrogen after menopause and other sites that synthesize estrogen become progressively crucial in the postmenopausal state. Given that the adrenal only generates low estrogen levels [5], it is evident that the bulk of postmenopausal women’s estrogen production occurs in periphery such as breast tissue. This non-ovarian in situ biosynthesis of estrogen could contribute to almost all of the total estrogen produced in postmenopausal women [4].

By binding and activating ER, estrogens facilitate the growth and survival of both benign and malignant breast epithelial cells. The activated receptors in turn act on to gene promoters in the nucleus and switches on many other genes required for cell division, cell death inhibition, de novo formation of blood vessels, and protease activity. With rapid proliferation, there is the potential for genetic errors and consequent predisposition to malignant cellular transformation [6]. Alternatively, estrogen metabolites may have core genotoxic effects that cause cellular transformation as well as DNA damage. Endogenous estrogen demonstrate its carcinogenic effects via metabolic activation [7,8].

Development and progression of hormone-responsive breast cancers can be interrupted via simply interfering with synthesis of estrogens. Aromatase is the main enzyme that catalyzes androgens’ transformation to estrogens which may result to a microenvironment with increased estrogen production in postmenopausal breast [9]. Thus, aromatase is one of the most important targets in the breast cancer treatment strategy for postmenopausal women. It has been nearly three decades since the third-generation aromatase inhibitors (AIs) were introduced and ascertained their superiority to ER blockers (tamoxifen) both in adjuvant setting and advanced disease [10–12]. Hereby, treatment with a third-generation AI has become the advised first-line endocrine therapy for ER-positive postmenopausal breast cancer cases [13]. However both de novo and acquired resistance may occur, which cause a limiting effect over AI therapy efficacy.

Along with tumor cells, non-cancerous cells comprising the tumor microenvironment (TME), which are known as TAS (e.g., adipocytes, fibroblasts and inflammatory cells), recognized as critical mediators of tumor progression (Figure 1). The supply of estrogen for postmenopausal breast tumors in both tumor and TAS cells emerges from circulatory uptake and local synthesis [14]. The significance of local synthesis versus circulatory uptake for the production of estrogen to cancer cells has been disputed, and while most breast carcinoma cells produce aromatase, most analyses have shown the significance of circulatory uptake of estrogen [14,15]. Treatment with AIs outcomes in complete suppression of the synthesis of whole body estrogen, but cancer cells have the ability to boost up the of local estrogen biosynthesis via crosstalk with TAS cells. We and other groups demonstrated the overexpression of aromatase in TME compared to breast tumor cells [16–18]. AI resistance mechanism may be different when the tumor cells are able to exploit the endogenous aromatase enzyme for local production of estrogen which will be eventually used in favor for the tumor progression. This emphasizes the clinical importance of local tissue aromatase activity and the need for a reliable method for its measurement. Therefore we investigated local specific aromatase activity levels both in postmenopausal IDC breast cancer cases biopsy specimens who did not received any endocrine therapy and healthy premenopausal health breast tissues.

**Fig. 1.**
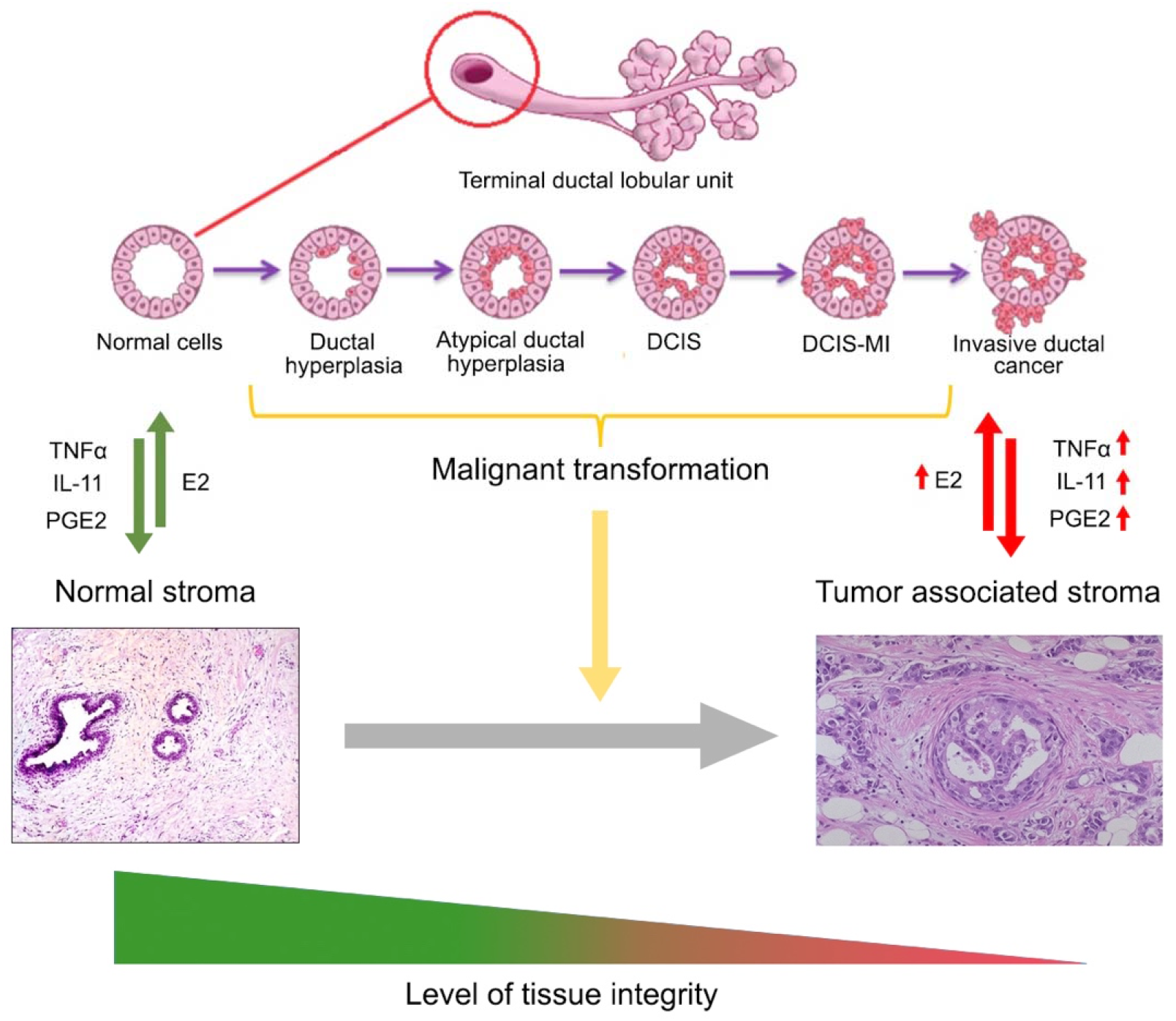
Crosstalk between stroma and tumor tissues during progression of IDC. In healthy conditions, the crosstalk maintains tissue equilibrium and integrity. During epithelial carcinogenesis, the efferent signaling changes. This leads to alterations in the stroma. Malignant epithelial cells enrich the population of adipose fibroblasts by secreting large amounts of cytokines such as tumor necrosis factor (TNF) and interleukin 11 (IL-11) to inhibit differentiation of preadipocytes into mature adipocytes; thus, causing a desmoplastic Furthermore, they secrete prostaglandin E2 (PGE2) and other unknown factors which leads increased production of aromatase at TAS tissue Thus, the new established crosstalk between tumor cells and tumor associated stromal cells leads to invasion and subsequently to metastasis.

Several techniques were used to assess the activity of aromatase. For demonstration of featured aromatase activity in clinical or in vivo studies a metabolic ratio of 17β-estradiol (E2) to testosterone is often used. Underlying reason for this application is the concentrations of substrates are not strictly controlled and the rate of formation can’t be directly measured with currently used techniques. Two biochemical assays are featured in most of the breast tissue aromatase measurement studies. In the first, also called ‘product identification assay’, aromatase activity is determined by measuring the formation of E2 from testosterone in homogenized tumor tissue [19,20]. In the other assay aromatase activity is determined by measurement of titrated water released in the conversion from titrated androstenedione to estrone (E1), also in homogenized tumor tissue [21]. The disadvantage of the latter assay is that the product of conversion by aromatase is not identified and thus possibly other processes in which tritium is released from androstenedione may influence the results. One of the purposes of current study was to develop a sensitive microsomal assay for measuring tissue aromatase activity and to show that this method can be used to measure activity in tumor and stromal regions of the postmenopausal breast with IDC.

## 2. Materials and methods

### 2.1. Patient selection

Tumors and nearby TAS tissue were acquired in the Department of Surgery at the Cerrahpasa University in January 2010-December 2012 from 20 women who received no adjuvant treatment before surgery and who received mastectomy or breast conservation surgery owing to invasive ductal breast disease. Surgical resected tissues were diagnosed and confirmed by a pathologic examination to correctly sample T and TAS tissue. Obtained samples were instantly frozen with liquid nitrogen and maintained at −80°C until used for the aromatase activity determination. All patients were classified as luminal A (ER(+), PR (+), HER2 (-) and postmenopausal. Tumor cells were reported as positive with prominent nuclear ER and PR immunostaining. In routine pathological interpretations, the ER and PR conditions of the patients were defined by immunohistochemistry on the formalin-fixed, paraffin-embedded sections of clinical specimens. Immunohistochemistry was performed using a rabbit monoclonal antihuman ER antibody (clone SP1; Thermo-Scientific, MA, USA) and a polyclonal rabbit antihuman PR antibody (clone 16, Novocastra, Leica Microsystem, Wetzlar, Germany). Two pathologists have assessed the immunohistochemical stainings. Over 10% of cell’s nucleuses stained were recognized as ER or PR positive. In addition chromogenic intensities of the receptors were assessed as weak and strong. Premenopausal women (age range = 20–40 years) were included in the study as the control group. A total of twelve healthy breast tissue samples (N) were collected during the reduction mammoplasty surgery. Prior to surgery, breast ultrasound was done and after operation, all samples were confirmed pathologically clean, thus none have a history of cancer and any metabolic disease known to them. Study protocol was approved by the Ethical Committee of Istanbul University School of Medicine (no: 2011/1808-804). Informed written consent about the study was obtained from each patient.

### 2.2. Microsome isolation

Tissues (100–150 mg) were pulverized with liquid nitrogen using a ceramic mortar and pestle and transferred to 1.5 mL micro centrifuge tubes containing 0.5mm diameter zirconium oxide beads (Next Advance, NY, USA) and homogenization buffer (1x PBS (pH 7.4), 20% glycerol, 0.1 mM dithiothreitol, 1 mM EDTA). 1 mL of homogenization buffer was used for each 100mg of tissue sample. Protease inhibitor cocktail (Abcam, MA, USA) was added just before homogenization. A bead beater homogenizer was used (SpeedMill Plus, Analytik Jena, Jena, Germany) for effective homogenization of breast tissues. Differential centrifugation technique was applied, centrifugation at 9000 g and 120000 g respectively, in order to obtain the microsomal fraction. A solubilization buffer was prepared (1x PBS (pH 7.4), 20% gliserol, 1 mmol/L EDTA, 1mmol/L sodium cholate and 0,2% Tween 20) for resuspension of the pellet. After storing the samples at 4°C overnight on a shaker, protein levels were detected by Pierce Bicinchoninic acid (BCA) protein assay (Thermo-Scientific, IL, USA). Microsomes were stored at −80°C until used.

### 2.3. Western blot analysis

From each microsomal fraction sample, 20 μg of proteins were resolved by 4–20% Mini-PROTEAN TGX precast gels (Bio-Rad Laboratories, CA, USA) and transferred to a PVDF membrane (Bio-Rad Laboratories, CA, USA). The membrane was first blocked in the blocking buffer (5% nonfat dry milk, 0.1% Tween-20; in PBS) for 1 h at room temperature and then Prior to incubation of the membrane with anti-aromatase antibody (1:200; Abcam, USA) in blocking buffer (5% nonfat dry milk, 0.1% Tween-20; in PBS) for overnight at 4°C, 1h of incubation was performed with the same buffer. The membrane was washed and then incubated with an HRP-conjugated goat anti-rabbit secondary antibody (1:3000; Abcam, USA) for 1 h at room temperature and detected by ChemiDoc MP system (Bio-Rad Laboratories, CA, USA) respectively. Blot stripping and β-actin antibody (1:1000; Santa Cruz Biotechnology, USA) reprobing were applied in order to confirm equal protein loading. All the gels and blots were analyzed via ImageLab software version 4.1 (Bio-Rad Laboratories, CA, USA).

### 2.4. Microsomal incubation assay

In a total reaction volume of 1 mL potassium phosphate buffer (0.5 M pH 7.4), the activity of microsomal fractions to catalyze the transformation of testosterone to E2 was determined. Testosterone (total concentration of reaction 0,5 μ g / mL) was added to the reaction solution and pre-incubated at 37°C for 5 min. By adding the NADPH regenerating reagents (NADP 0.4 mM, glucose-6-phosphate (G-6-P) 4 mM, G-6-P dehydrogenase 1 U/mL, and 2 mM MgCl_2_), the reaction was launched and incubated at the same temperature for 20 minutes. The process was ended by adding a 10% (w / v) trichloroacetic acid to 100 μL and put on ice. As an internal standard, E1, 0.5 μL of 100μg / mL solution, was loaded into each tube. Furthermore, a blank sample with the same reagents and conditions was prepared, but without substrate.

### 2.5. Estradiol extraction

The extraction of the produced E2 was performed via a modified version of solid phase extraction (SPE) protocol which was described by Newman et al [22] previously. A 20-place vacuum manifold (Vac Elut 20, Agilent Technologies) was employed during the whole SPE process. All solvents were LC-MS (liquid chromatography-mass spectrometry) grade. Briefly, ethanol was used to prime end-capped columns (Sep-Pak C18 Plus Short Cartridge, Waters), followed by preparation of the sorbent for sample loading with water. Samples were passed through columns for sample loading and interfering polar substances were washed out by water. E2 was eluted from columns with 1.5 mL of methanol (MeOH) into microfuge tubes. Eluates were dried at 40°C under a steady stream of medical-grade N_2_ and resuspended with 100 μl MeOH then transferred to glass vials with inserts for LC-MS/MS analysis.

### 2.6. Monitorization of estradiol formation

For chromatographic separation and analysis, Agilent 1200 Series HPLC (high pressure liquid chromatography) system (Agilent Technologies, USA) was used with a C18 column (3.0×30mm; 1.8μm, Zorbax SB-C18) including an online filter. Temperature of the column was kept at 40°C. Acetonitrile (A) and 0.1% formic acid in water (B) were used as mobile phases. The initial mixture ratio of A and B and the flow rate of the mobile phase were 90:10 and 0.6 ml/min, respectively. After 3 min mobile phase B was increased to 40% for 1 min. Then a linear return to initial conditions was performed over 1.5 min. The injection volumes were 20 µL and run time for per sample was 5.5 min.

Mass spectrometric analysis of analyte formation was performed using an Agilent 6460 Triple Quad System (Agilent Technologies, USA) mass spectrometer coupled with Electrospray Ion Source with Agilent Jet Stream operated in positive multiple reaction monitoring (MRM) mode with unit resolutions set at 401 and 383 at Q1 and Q3 respectively. Monitoring of the m/z transitions were monitored for the quantification of E2 levels in samples. The retention time for E2 was 0.9 min and 1.14 min for testosterone. Particular fragment ions were selected to pass through Q3. These were selected for quantitation and confirmation. For E2, the product ion at m/z 158.9 is more intense than the product ion at m/z 133. Therefore, the MRM transition 255.1 to 158.9 was used for quantitation and the m/z 133 ion was considered as qualifier ion. For testosterone the MRM transition 289.1 to 96.9 was used for quantitation and the m/z 108.9 ion was considered as qualifier ion (Figure 2). Optimized parameters for source were as follows: gas temperature = 350°C, gas flow = 13 l/min, nebulizer = 40 psi, sheath gas heater = 400°C, sheath gas flow = 12 l/min, capillary voltage = 6000V, noozle voltage = 2000V. Scan time was set at 0.01 s and collision gas pressure was set at 1.5mTorr. The data were collected and quantified using Agilent Mass Hunter software version 2.0.6 (Agilent Technologies, USA).

**Fig. 2.**
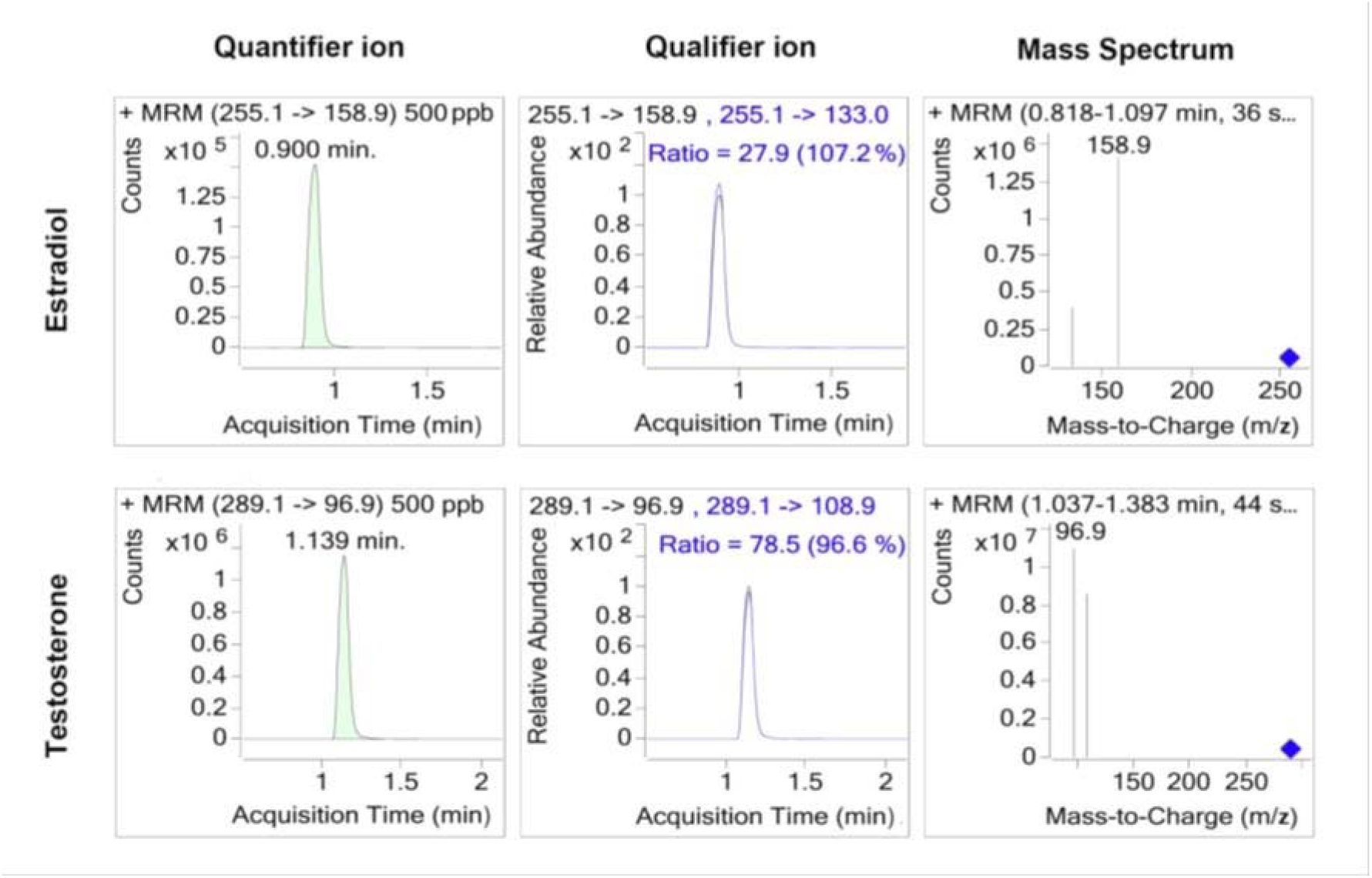
MRM mass spectrums of the quantifier and qualifier ions for estradiol and testosterone.

### 2.7. Aromatase activity calculation

The calibration curve and study group samples were prepared and extracted. Standard curve ranging from 1 pg/μl to 1000 pg/μl was prepared for E2. MeOH was used to produce a stock solution for E2, E1 and testosterone of 1 mg/ml in order to prepare a series of working solutions. Calibrators contained E2 at concentrations of 1, 10, 100, 250, 500, 1000 pg/μl, quality controls (QCs) were 1 pg/μl, 20 pg/μl and 50 pg/μl at a volume of 500 μl. Area under the curve (AUC) values are used to calculate the concentration of E2 (pg/μl) in the unknowns samples. Aromatase activity is calculated as unit activity and presented as specific activity (U.mg^−1^) for each sample. Negative control samples did not receive microsome for the reaction mixture. Diluted microsomes from placenta tissue were used as a positive control. Total protein in patient samples quantified colorimetrically via BCA Protein Assay (Thermo Scientific, Pierce, USA). Assay was conducted according to manufacturer’s instructions.

### 2.8. Statistical analysis

All calculations were performed using SPSS Statistical Program version 22.0 (SPSS Inc. Chicago, IL, USA). The correlation between aromatase activity and patient characteristics was analyzed using the Goodman and Kruskal’s gamma test (G). The significance of differences in activity levels between tissue groups was determined by the Wilcoxon signed-rank test (WSR) or Mannwhitney U test (MU) as needed. ANOVA was performed for evaluation of the aromatase activity alterations together with the stage, grade and receptor status of the tumor. Two-sided tests were used to obtain all reported p-values and the results below 0.05 were considered as statistically significant.

## 3. Results

### 3.1. Demographic data and patient characteristics

In our study, 20 female IDC patients and 12 female breast reduction surgery patients were included. Patient characteristics for IDC patients are shown in Table 1. The mean age (± SD) and body mass index of the study group (n = 20) were 59.65± 11.94 years and 27.871 ±5.06 kg/m^2^ respectively.

**Table 1:**
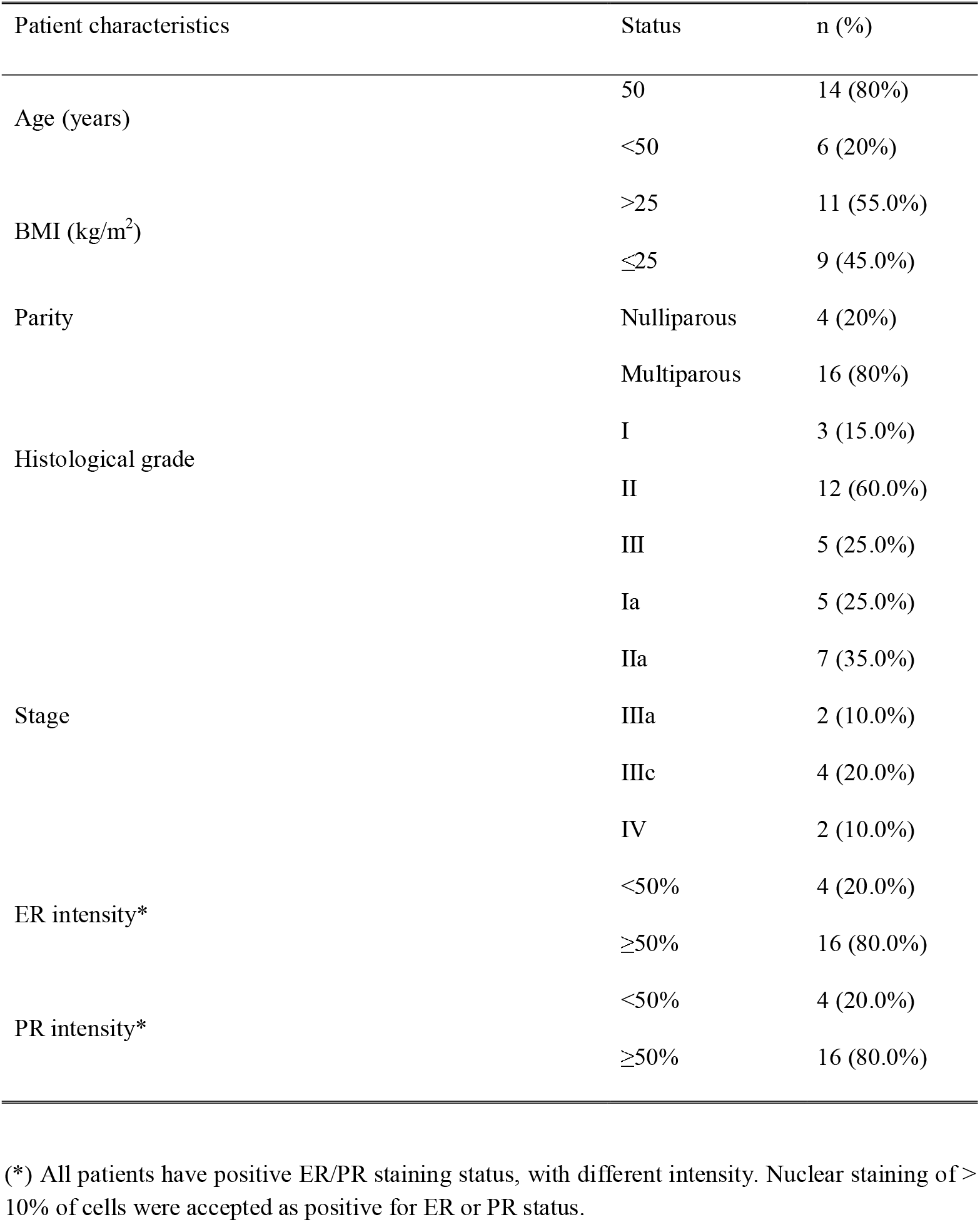
Characteristics of IDC patients (n = 20).

We investigated the possible associations between aromatase activity levels and patient characteristics (Table 2). Known breast cancer risk factors being over 50 years old and nulliparous were found to be correlated for patients who have higher aromatase activity within their TAS tissue compared to its tumor.

**Table 2:** Patient characteristics correlated with two fold and higher/lower difference between tumor and its paired TAS tissue.

| Patient characteristics | Status | Specific aromatase activity |  | p value |
| --- | --- | --- | --- | --- |
| | | TAS/T $\geq 2$<br>n (%) | TAS/T $< 2$<br>n (%) | |
| Age (years) | 50 | 2 | 4 | 0.027 |
|  | <50 | 12 | 2 |  |
| Parity | Nulliparous | 4 | 0 | 0.032 |
|  | Multiparous | 10 | 6 |  |
| PR intensity | <50% | 4 | 0 | 0.032 |
| | $\geq 50\%$ | 10 | 6 | |
| ER & PR intensity | ER<PR | 5 | 3 | 0.556 |
| | ER≥PR | 9 | 3 | |

### 3.2. Highest aromatase activities in TAS tissues

Overall, highest specific activity was observed in the patients’ TAS tissue. Among the patients approximately 3.5 fold increased activity was observed. The calculated activity of aromatase was higher in TAS breast tissue of 70% of the patients compared to their tumor tissue and the difference was statistically significant (Z = −2.85, p = 0.004, WSR). Specific aromatase activities of premenopausal healthy subjects’ breast tissue were found nearly 3 fold higher compared to tumor tissue level (p< 0.001, MU) (Figure 3). Aromatase expression levels were also checked with western blotting. Obtained results were in concordance with activity calculations. The expected 52-kDa aromatase was detected in all patients, and the amount of aromatase protein was greater in TAS tissue than the tumor tissue (Figure 4).

**Fig. 3.**
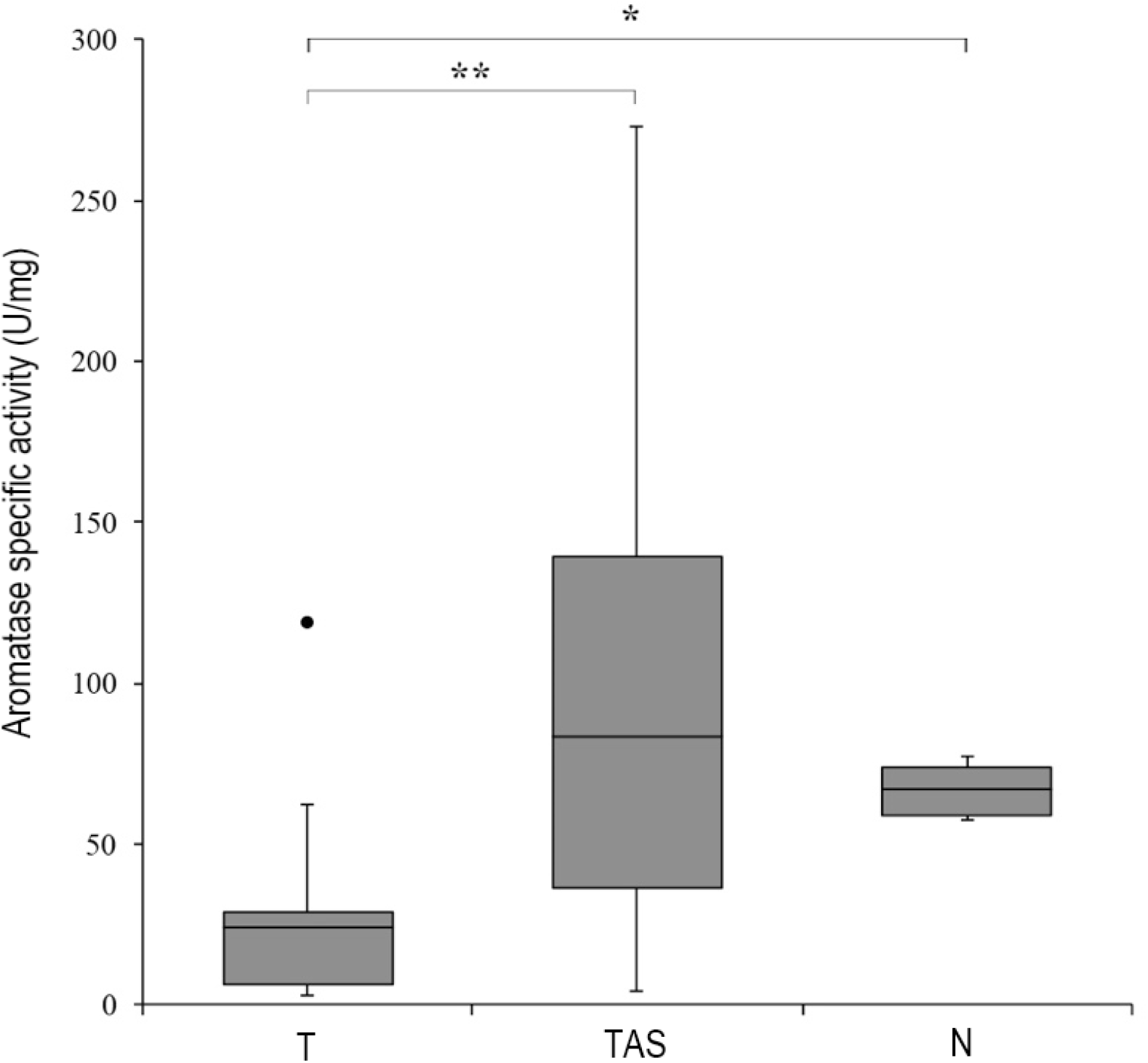
Specific aromatase activity levels among tissue samples. *Mann–Whitney U = 18, p < 0.001 two-tailed. **Wilcoxon signed-rank test: Z = −3.17, p = 0.004 two tailed. (•) refers to outlier.

**Fig. 4.**
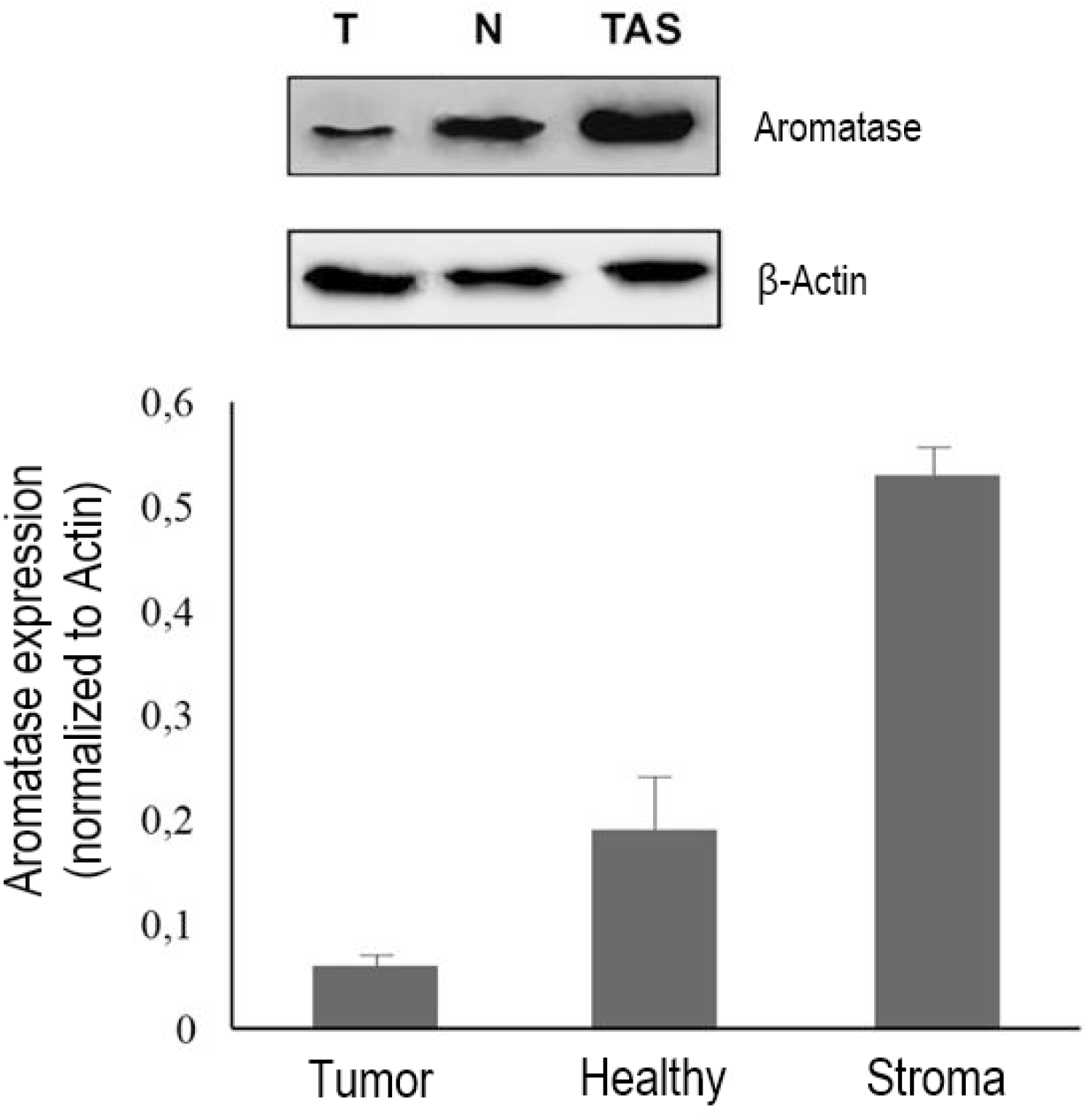
Western blot analysis of Aromatase in microsomal fractions of tumor, tumor associated stroma and healthy breast tissues. T: Tumor, TAS: Tumor associated stroma (TAS), N: Healthy breast tissues. Immunoblot signals were quantified by densitometry, and normalized with β-actin.

### 3.3. Grade and stage of tumor effects local aromatase activity

Most patients in the study group had disease of histological grade II or higher (17/20, 85.0 %), and stage of the disease is IIa or worse (15/20, 75.0 %). Regarding to the histological grade, both peripheral and tumor cells seem to have an input in estrogen production via aromatase (Figure 5). A notable change in tumor cells is observed which they are more actively produce estrogen in early phases of the disease (ANOVA Bonferoni test, p = 0.039 and p = 0.043, specific aromatase activity in Grade I compared to Grade II and III respectively) Aromatase activity levels in tumor tissue were ordered Grade I> Grade II> Grade III. Although there was no statistical significant difference for aromatase activity among tumor tissues when grouped according to their stages (p = 0.342), a slightly decreasing pattern was seen while the stage of the tumor increases. On the contrary aromatase activity is found to be increasing at TAS tissues (Figure 6).

**Fig. 5.**
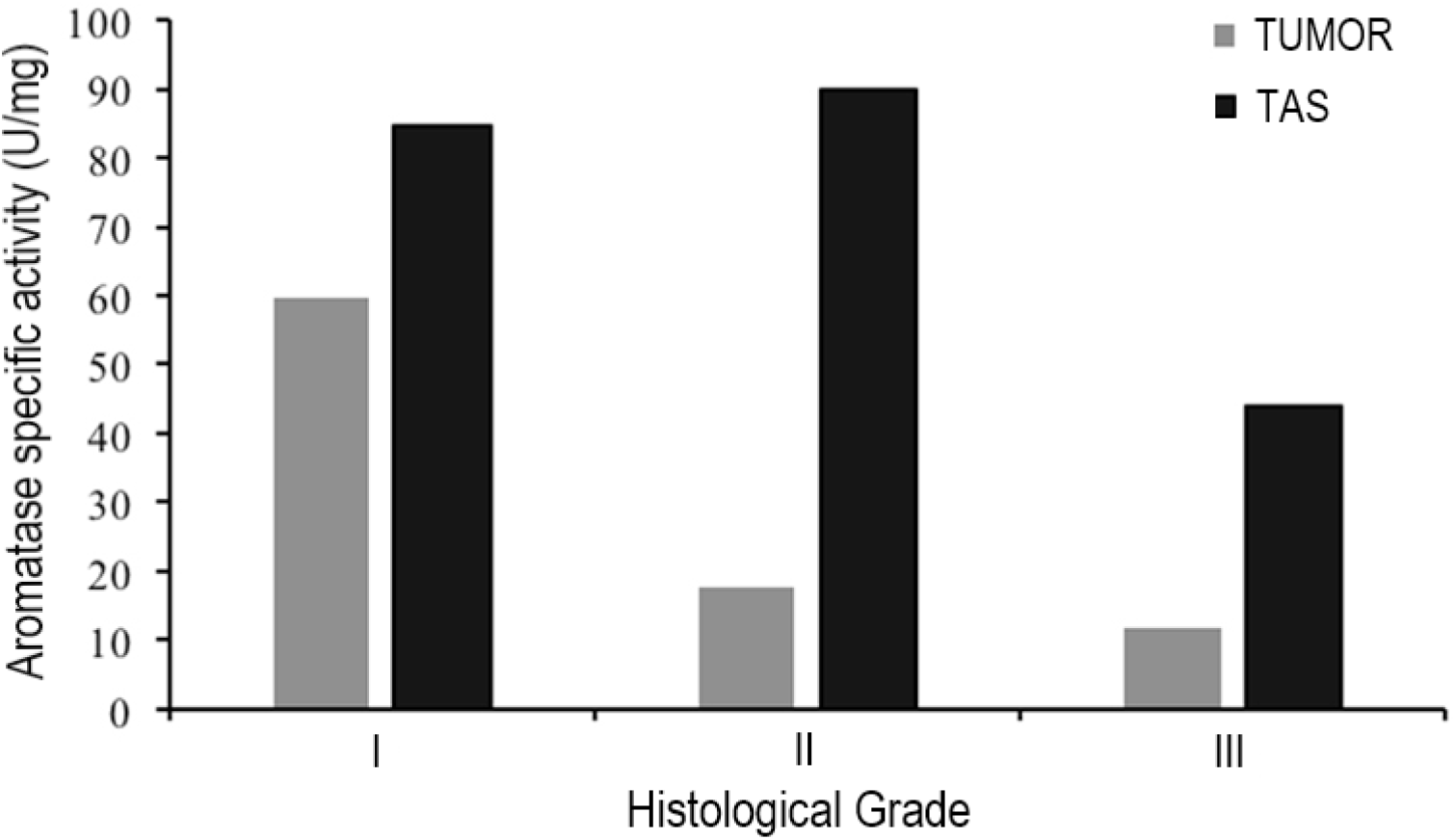
Aromatase activity levels among tissue groups according to the tumor histological grades.

**Fig. 6.**
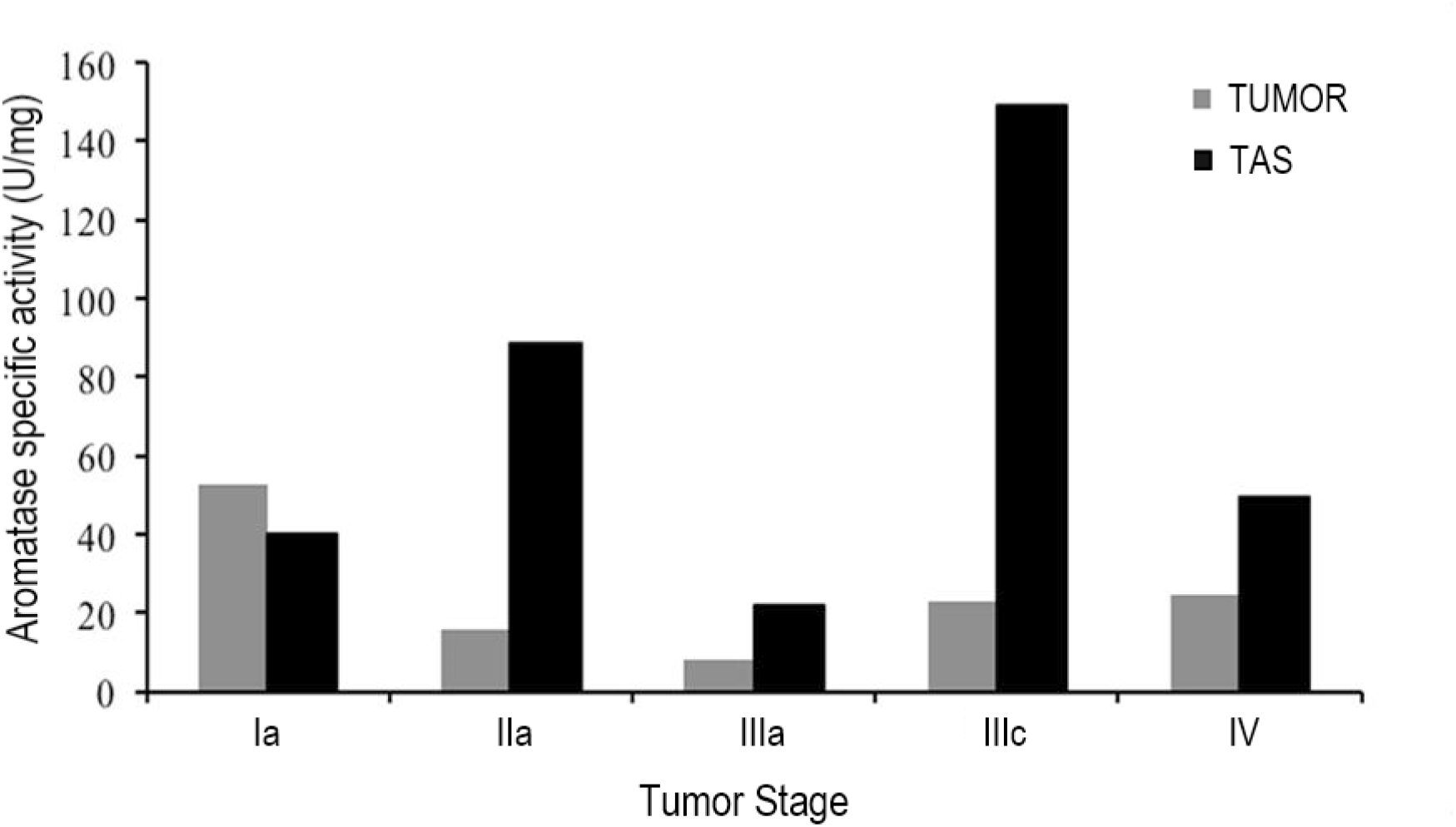
Aromatase activity levels among tissue groups according to the tumor stages.

### 3.4. High PR intensity decreases TAS tissue aromatase activity

All patients in the study group were ER/PR positive and have intensity levels over 10%. We grouped the patients according to ER/PR intensity levels (≥50% and < 50) of their tumor samples (Table 2). Higher PR intensity was correlated with cases which has two fold lower aromatase activity in TAS tissue compared to tumor tissue (p = 0.032, G). We did not find any significant correlation with ER intensity alone (data not shown) or both receptors together and the patient aromatase activity levels (p = 0.556, G).

## 4. Discussion

Estrogens are essential for the normal breast development and maintenance. Regrettably, because of their capacity to encourage cell proliferation, estrogens can also be damaging, which can also increase the risk of a woman developing breast cancer [23].

Breast cancer is exclusive in that it develops and prospers in stromal tissue, which provides structural and paracrine support for the growing tumor. As expected, the main fuel for this growth is the estrogen for the ER (+) IDCs. The most important source of estrogen in postmenopausal women that support the development and growth of breast cancers is the increased local production within the tumor tissue or in the surrounding stroma caused by aromatase overexpression [24]. It acts at the site of synthesis in a paracrine and/or intracrine manner. The importance of local synthesis versus circulatory uptake for supply of estrogen to the cancer cells has been debated and although most breast tumor cells express aromatase, most studies point towards a major importance of uptake of circulatory estrogen [15]. Studies have demonstrated that tissue estrogen levels were locally amplified however circulating levels were not and look as if to utilize hyperplastic effects on the mammary tissue in a paracrine manner [25,26]. Our data supports these findings where it points out main estrogen production site is the peripheral stromal tissue (TAS) in postmenopausal IDC cases (especially when the grade of tumor is high) and almost has the same aromatase activity level with premenopausal healthy breast tissue.

Ongoing research has shown that tumor progression depends not only on the specific characteristics of cancer cells but also on more complex interactions between tumor and host cells [27]. Recent evidence suggests that the tumor stroma profoundly influences tumor growth, angiogenesis and dissemination. TAS cells are believed to encourage tumorigenesis through various processes including remodeling of the extracellular matrix, suppression of immune response, and changes in stromal regulatory pathways influencing cancer cells’ motility and aggressiveness [28]. Although the importance of cancer-associated stromal cells in cancer progression is known, its relationship with other components of the tumor microenvironment has not yet been fully defined.

Earlier studies demonstrated that estrogens are able to active many signaling pathways in a rapid fashion via binding ERs in or near the plasma membrane. It has been more than a decade Nemere et al. coined the terms according to the location of estrogen action (NISS: nuclear-initiated steroid signaling, MISS: membrane-initiated steroid signaling) [29]. It is also well known that ER and PR expression levels have an effect on the response to endocrine therapy. Cases expressing only ER are tend to respond poorly to selective ER modulator (SERM) drugs, such as tamoxifen, compared to the ones with both receptors are expressed. The predictive value of PR expression has long been attributed to the dependence of PR expression on ER activity, with the absence of the PR reflecting a non-functional ER. The study conducted by Cui et al. demonstrated that PR expression is inhibited in breast cancer cells via PI3K/Akt/mTOR pathway, not via a reduction in ER levels or activity [30]. Furthermore, it was shown that estrogen stimulates association between ER and IGF-IR (via MISS ER activity), and this results in activation of IGF-I signaling via ERK1/2 [31]. An additional explanation came from a recently published preclinical data which suggests that PR activity can change where ER binds to DNA, directly modulating ER function and thereby potentially improving the tumor response to ER antagonists [32]. Here we hypothesized a possible molecular pathway according to ER/PR expression levels in tumor tissues (Figure 7). Our results showed us the increase of aromatase activity in TAS tissue negatively correlates with the expression of PR, independent from ER expression level in tumor cells. A logical explanation of this phenomena may be the imbalance of MISS and NISS ER activity in tumor cells, that are induced by high levels of E2 in tumor microenvironment, provided by TAS cells. GF signaling may also have an effect on the imbalance for the favor of NISS which may support the development of endocrine therapy resistance (poor prognosis). Therefore, high aromatase activity in tumor neighboring tissue specimens may serve as an upstream indicator for such cases. However the underlying mechanisms are still largely unknown and further investigations are required with larger study groups to solidify our findings.

**Fig. 7.**
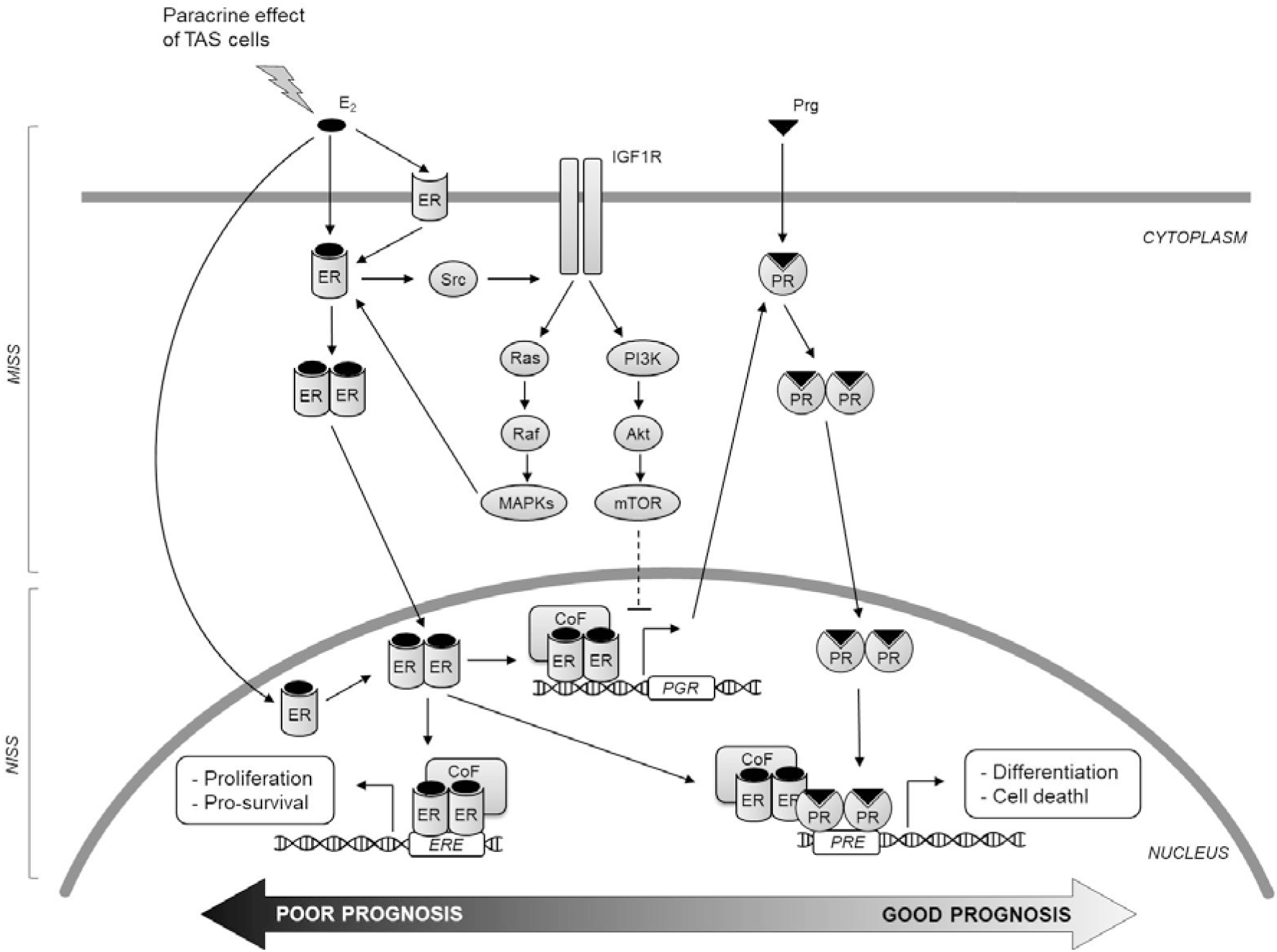
The hypothesized molecular mechanism underlying endocrine therapy resistance induced by high E2 levels and PR expression alteration.

SERMs and AIs were developed for inhibiting ER activation and stopping breast cancer growth [33]. Currently two possible options are available for the adjuvant endocrine therapy of postmenopausal women: 5 to 10 years of tamoxifen or 5 to 10 years of an AI. The therapy of high-risk patients, who have tolerated endocrine therapy well for the first 5 years, can be extended further than the first 5 years [34]. According to the disease free survival outcome superiority of AIs compared to tamoxifen [35,36], switching strategies may also be applied [37]. But still, recurrence within 10 years and eventually progress to incurable metastatic disease can be seen among 20% of the patients [12]. The molecular mechanism behind how some ER (+) breast cancer tumors develop resistance to the hormone drugs tamoxifen and aromatase inhibitors is still unknown. Not only the microenvironment of the tumor, the ability of carcinoma cells to utilize the endogenous aromatase enzyme for estrogen supply is also important, regarding the AI resistance. Magnani et. al [45] observed that AI treatment itself causes aromatase gene (*CYP19A1*) amplification that promotes local autocrine estrogen signaling in AI-resistant metastatic patients. Furthermore, our data tempts us to suggest that in early stages of the cancer, tumor cells promote the local autocrine estrogen signaling even before taking any endocrine therapy. We also observed that in cases with high stages the aromatase activity is higher in TAS tissue which points out that the production site of E2 is shifted. The possible reason for this may be the necessity to establish certain factors, such as TNF-alpha and IL-11, expression in order to modulate the aromatase expression in stromal tissue.

It appears the in-house LC-MS/MS method is applicable for routine measurement in clinical laboratories. Addition to its high selectivity and accuracy, the other advantage of the method is that the analytes (E2 and testosterone) do not require derivatization prior to measurement, which makes it fast and simple to apply. Liquid chromatography-mass spectrometry systems with ESI technology have become essential for clinical biochemistry laboratories within last decade [38]. Thus the method for aromatase activity measurement proposed here has the potential to be used as a clinical test, after the completion of further studies for standardization according to the bioanalytical guidelines.

We recognize that there are certain constraints to this research. Regardless of the small amount of cases, to eliminate this restriction, suitable analysis was performed. Due to breast-conserving surgery applications, the major challenge was to obtain appropriate quantities of breast tissue samples coupled as tumor and nearby stromal tissues. We were also obliged to exclude the patients from the study, who had adjuvant chemotherapy prior to surgery, in order to eliminate the additional effects on the aromatase activity levels to be observed. Our control group is consisting of premenopausal healthy women patients, applied to our clinic for breast reduction surgery. Due to ethical reasons no healthy older patient was asked for a clinically unnecessary breast biopsy, therefore postmenopausal healthy tissue samples were unavailable.

## 5. Conclusions

Detecting the breast cancer early and finding the effective therapy for it in a personalized manner are equally important. Our endgame will be to establish an accurate molecular profiling of patients with estrogen driven breast cancer where we can help physicians to enhance current treatment strategies in a more personalized way. Comparative measurement of aromatase activity levels can be a good starting point, nevertheless, evaluation of other key enzyme activities in steroidogenesis pathway that effect local estrogen levels should be considered for future studies. The proof of concept methodology presented here can be modified to be used for measuring other relevant steroid targets.

## Data Availability

The data that support the findings of this study are available from the corresponding author upon reasonable request.

## List of abbreviations

AI: Aromatase inhibitor
Akt: Protein kinase B
AUC: Area under the curve
E1: Estrone
E2: 17-beta estradiol
EDTA: Ethylenediaminetetraacetic acid
ER: Estrogen receptor
ERK: Extracellular signal-regulated kinase
G-6-P: Glucose-6-phosphate
GF: Growth Factor
HER2: Human epidermal growth factor receptor 2
HPLC: High pressure liquid chromatography
IDC: Invasive ductal carcinoma
IGF-IR: Insulin-like growth factor 1 receptor
IL-11: Interleukin-11
LC: Liquid chromatography
MeOH: Methanol
MISS: Membrane-initiated steroid signaling
MRM: Multiple reaction monitoring
MS: Mass spectrometry
mTOR: Mammalian target of rapamycin
MU: Mannwhitney U test
N: Healthy breast tissue
NISS: Nuclear-initiated steroid signaling
PBS: Phosphate buffered saline
PI3K: Phosphoinositide 3-kinase
PR: Progesterone receptor
SERM: Selective ER modulator
SPE: Solid phase extraction
T: Tumor
TAS: Tumor-associated stroma
TME: Tumor microenvironment
TNF-alpha: Tumor necrosis factor alpha
WSR: Wilcoxon signed-rank test

## Acknowledgements

We are grateful to Saffet Çelik from Trakya University for his expert technical support with mass spectrometry analysis.

## Statement of Author Contributions

Mete Bora Tüzüner: Conceptualization, Project administration, Writing –Original draft preperation, Methodology. Tülin Öztürk: Investigation, Resources, Writing – Original draft preparation. Sennur Şlvan: Investigation, Resources. Hande Turna: Resources. Türkan Yurdun: Formal analysis. Hülya Yılmaz Aydoğan: Investigation, Formal analysis, Visualization. O uz Öztürk: Project administration, Supervision.

## Compliance with Ethical Standards

### Ethics Approval and Consent to Participate

All women provided a signed informed consent. The study was approved by the Ethical Committee of Istanbul University School of Medicine and with the 1964 Helsinki declaration. All procedures were followed in accordance with the ethical standards of the responsible committee on human experimentation (The Scientific Research Projects Ethical Board of Istanbul University School of Medicine, no: 2011/1808-804).

